# Anomalies in regional and chronological distributions of Omicron BA.1.1 lineage in the United States

**DOI:** 10.1101/2024.08.14.24311991

**Authors:** Hideki Kakeya

## Abstract

This study compares the collection dates and locations of the Omicron BA.1 lineage and other major SARS-CoV- 2 mutants registered in NCBI GenBank and provides a detailed analysis of the emergence patterns of pure reverse mutants, which contain only reverse mutations and no other mutations in the surface glycoprotein. The results indicate that Omicron BA.1.1 and its pure reverse mutants were widely distributed throughout the United States from the early days of their emergence, showing a statistically significant difference compared to other major variants, which spread from a small number of sources. The peak emergence of BA.1.1 and BA.1.1.18 pure reverse mutants occurred a few weeks before the peak of all collected samples, whereas the peak of pure reverse mutants in major BA.1 variants and BA.2 coincides with the overall sampling peak. Although the peaks of BA.1.x collections are not all synchronous, the peaks of pure reverse mutants in the BA.1 lineage completely overlap, with the number of such mutants declining abruptly after the peak. These regional and temporal anomalies in the Omicron BA.1 lineage, especially in the BA.1.1 lineage, are virtually impossible to explain by current theories of natural mutation and spread by human-to-human infection.

## Introduction

Many studies have tracked local, national, or world-wide mutations and transmissions of SARS-CoV-2 and its variants. Regarding the original Wuhan strain, Pekar et al. [1] and Worobey et al. [2] claimed SARS-CoV- 2 had originated in the Huanan Seafood Market, based on early mutations and sampling locations at the onset of the outbreak, while allegations of sampling bias were raised against them in subsequent publications [3,4]. Zhan et al. compared early mutations of SARS-CoV and SARS-CoV-2 to find that the latter was well adapted to human transmission from the early days of its emergence, which is inconsistent with a hypothesis that the virus had a zoonotic origin [5].

The first major mutation observed in SARS-CoV-2 was D614G in the surface glycoprotein (spike protein), whose spreads in the United Kingdom (UK) [6] and the world [7] were investigated in previous studies. It is known that D614 is unstable in vivo, not only in humans but also in hamsters [8]. Therefore, the emergence of D614G mutation through human-to-human transmission in the earliest days of the pandemic is quite natural.

After the D614G mutation, many other mutations followed, leading to the emergence of major variants of SARS-CoV-2. Among them, B.1.1.7 (Alpha variant), which is believed to have originated in the UK, and B.1.617.2 (Delta variant), which is believed to have emerged in India, prevailed globally from the late 2020 to the mid-2021. The dynamics of Delta variant transmission in the UK were analyzed in detail [9], where one or a few epicenters were visible for each mutant.

Most mutations among the major variants of concerns (VOCs) are independent of one another, with high dN/dS ratios [10,11] in the spike protein consistent across all VOCs [12]. Hassan et al. surveyed mutations in various VOCs across continents to find that many mutations were specific to each location [13], which could have contributed to the emergence of independent mutations.

Among the major VOCs of SARS-CoV-2, the Omicron variant, which emerged in November, 2021, is significantly different from other VOCs. The Omicron variant includes 30 or more non-synonymous (N) mutations and only one synonymous (S) mutation in the spike protein [14], whereas previous VOCs had only around 10 spike mutations. Phylogenetic analysis indicates that the Omicron variant did not emerge from the other precedent VOCs [15].

Initially, the origin of the Omicron variant was considered to be either unknown human population under strong selective pressure to escape vaccine-induced immune response, incubation in an immunocompromised patient, or evolution in a non-human host before spilling over back to human [16]. However, highly vaccinated populations in developed countries are well-monitored by health administration, making unnoticed community spread practically impossible. Regarding chronical immune escape in a single human subject, reported counts of mutations in immunocompromised patients are around 10 or fewer [17–19], which is not comparable to the number observed in the Omicron variant.

Regarding evolution in a non-human host, Wei et al. suggested that the Omicron variant evolved in mice [20], which was later supported by Zhang et al. [21]. It is known, however, that the original strain of SARS-CoV- 2 does not infect mice [22]. Kakeya et al. indicated that a lab origin of the Omicron variant is likely [23], possibly caused by a spill-over from transgenic mice [24].

Tanaka and Miyazawa found that the Omicron variants BA.1, BA.1.1, and BA.2 registered in NCBI (National Center for Biotechnology Information) GenBank comprise sequences with a single reverse mutation and no other mutations in the spike protein [25]. Kakeya and Matsumoto observed reversions in D614G of the spike proteins in the Delta variant (B.1.617.2) and the Omicron variant BA.2 in the late stages of their community spread [26]. Following these studies, Kakeya and Kanazaki compared the histograms of mutations in the spike proteins of major variants, where BA.1 is outstanding in its high reverse mutation rate and low variety of mutations [27].

The epidemiology of the Omicron variants has been investigated in many countries. Though some studies, such as those in Chile [28] and Taiwan [29], analyzed only chronological changes, many studies surveyed both chronological and geographical transitions of infection. In Brazil, several studies have been conducted locally and nationally. One study found that Omicron variant was mostly imported through the state of São Paulo, which dispersed the lineages to other states and regions of Brazil [30]. Another study examined the spatiotemporal dispersion of emerging SARS-CoV-2 lineages, including Omicron variants, from Sergipe State, which is located in northeastern Brazil, to find that highly populated cities acted as epicenters of circulation for each lineage regardless of vaccination status [31]. Another study performed SARS-CoV-2 genomic surveillance in the southeastern region of São Paulo State to observe the regional emergence of the BA.2 lineage during its initial dissemination [32].

In Mexico, the surge of Omicron BA.1 in the end of 2021 began at the east end of the country and spread westward [33]. In the UK, Omicron BA.1 was confirmed to have spread from London [34], which also holds true for Omicron BA.2 [35]. In Hong Kong, the spatiotemporal pattern of Omicron BA.1, BA.2, and Delta AY.127 were analyzed using phylogenetic trees and mathematical modeling of transmission [36]. In South Africa, Omicron BA.4 and BA.5 were confirmed to have spread from Johannesburg and Durban, respectively [37]. In Cyprus, transmissions of Omicron BA.1, BA.2, and BA.5 between Cyprus and other countries were traced [38]. As these studies demonstrate, spread of emergent variants is generally traceable.

Surprisingly, despite the numerous studies on the spread of Omicron variants worldwide, few studies have been performed to track the spread of the Omicron variants in the United States (US). While one study surveyed attack rates of various Omicron variants state by state, it did not provide chronological change in each state [39]. The author has recently investigated the spread of Omicron BA.1 and found a wide prevalence of reversion mutants from the beginning of the emergence, with no clear epicenters identified for most mutants [40].

This paper investigates how major SARS-CoV-2 lineages, including Alpha, Delta, BA.1, and BA.2, spread in the US. Specifically, Omicron BA.1.1, which has the largest number of registrations in GenBank of all the SARS-CoV-2 lineages, and its reverse mutants are analyzed in detail.

## Methods

The data of SARS-CoV-2 registered in GenBank were accessed June 2023 through July 2024. For each sequence, the accession number, submitter name, date of registration, country of collection, state (if collected in the US), and date of collection were gathered. The analysis focused on lineages prior to Omicron with more than 40,000 entries (B.1, B.1.1.7, B.1.2, B.1.617.2, AY.3, AY.25, AY.25.1, AY.39, AY.44, AY.100, AY.103), Omicron BA.1 variants with more than 20,000 entries (BA.1, BA.1.1, BA.1.15, BA.1.18, BA.1.20, BA.1.1.18), and other Omicron variants with more than 100,000 entries (BA.2, BA.2.12.1).

Data sampled in the US with their collection dates and locations (names of the states) registered were analyzed, where heatmaps of the first 100 samples for each variant were generated and compared. Data with the collection dates significantly prior to the confirmation of each variant, deemed as data errors, were removed from the analysis. When collection dates were the same, earlier registrations were counted as earlier samples. The total count of sampled collections in the top three and the top five states in the US were compared among different lineages.

Spike protein sequences of BA.1.1 including only reverse mutations, referred to as pure reversion mutants (PRM), were retrieved from GenBank, where the dates and locations of collection were also retrieved for each PRM. Data with missing reads were included in the analysis except for those including missing reads at the mutation points of Omicron BA.1.1, where missing amino acids were assumed to be the same as those of the consensus sequence.

The date and location data of BA.1 derivatives with more than 200 and fewer than 2000 registered spike protein sequences (BA.1.1.7, BA.1.1.8, BA.1.1.10, BA.1.1.15, BA.1.1.16, BA.1.1.17, BA.1.13, BA.1.14, BA.1.14.1, BA.1.15.1, BA.1.19, BA.1.21) were also downloaded from GenBank in May 2024 for comparison. Data of variants comprising more than 90 entries with collection dates and locations (ECDL) in the US, were used as a control group.

US data with collection dates and locations (state names) were analyzed, counting the number of states where the first 20 samples (SF20) were collected for each PRM pattern with more than 100 entries. When collection dates were the same, earlier registrations were counted as earlier samples here also. Statistical tests were conducted between the numbers of SF20 in the PRM group and the control group. PRM patterns with more than 50 registrations by December 27, 2021 (the first weekday after Christmas 2021) were analyzed, and the locations (states) of collection were expressed as heatmaps for each PRM, using the first 100 samples for mapping when the count of samples exceeded 100 by the above date.

To evaluate the reliability of data registered in GenBank, the rate of complete reads at Omicron mutation points of the spike protein and the ratio of organizations submitting ECDL for each lineage were calculated. To see the difference between the data registered in GenBank and GISAID (the Global Initiative on Sharing All Influenza Data), the ratio of entries including all the spike mutations (EIASM) specific to the consensus sequence of Omicron BA.1 variant (EIASM-BA.1) was calculated for the data registered to GISAID worldwide, in the US, and by the Centers for Disease Control and Prevention (CDC) of the US. Weekly ratios of EIASM-BA.1 in the end of 2021 and the beginning of 2022 were also calculated to assess read stability during the early days of the Omicron surge. The temporal changes in the amount of PRM and all sequences registered to GenBank by organizations other than the CDC were also analyzed to see the consistency of data registered by different organizations.

## Results

The locations (state of the US) where the first 100 samples were collected are expressed with heatmaps for the variants preceding the Omicron variant that have more than 40,000 spike protein sequence registrations, Omicron BA.1 derivatives with more than 20,000 spike protein sequence registrations, and other Omicron variants with more than 100,000 spike protein sequence registrations, as shown in Figure 1. The lineage B.1, which has more than 40,000 registered sequences, was excluded from the analysis because it took seven months for the number of ECDL in the US to reach 100. Table 1 lists the sum of sample counts in the top three/five states and the dates of the first and the 100th collection for each variant. As this table shows, Omicron BA.1.1 lacks clear epicenters, with samples collected evenly across many states. Under the normal distribution based on the data of 18 variants listed in Table 1, Omicron BA.1.1 has significantly smaller counts of samples for both the sum of top three states (*p* = 0.027) and top five states (*p* = 0.020).

**Figure 1.**
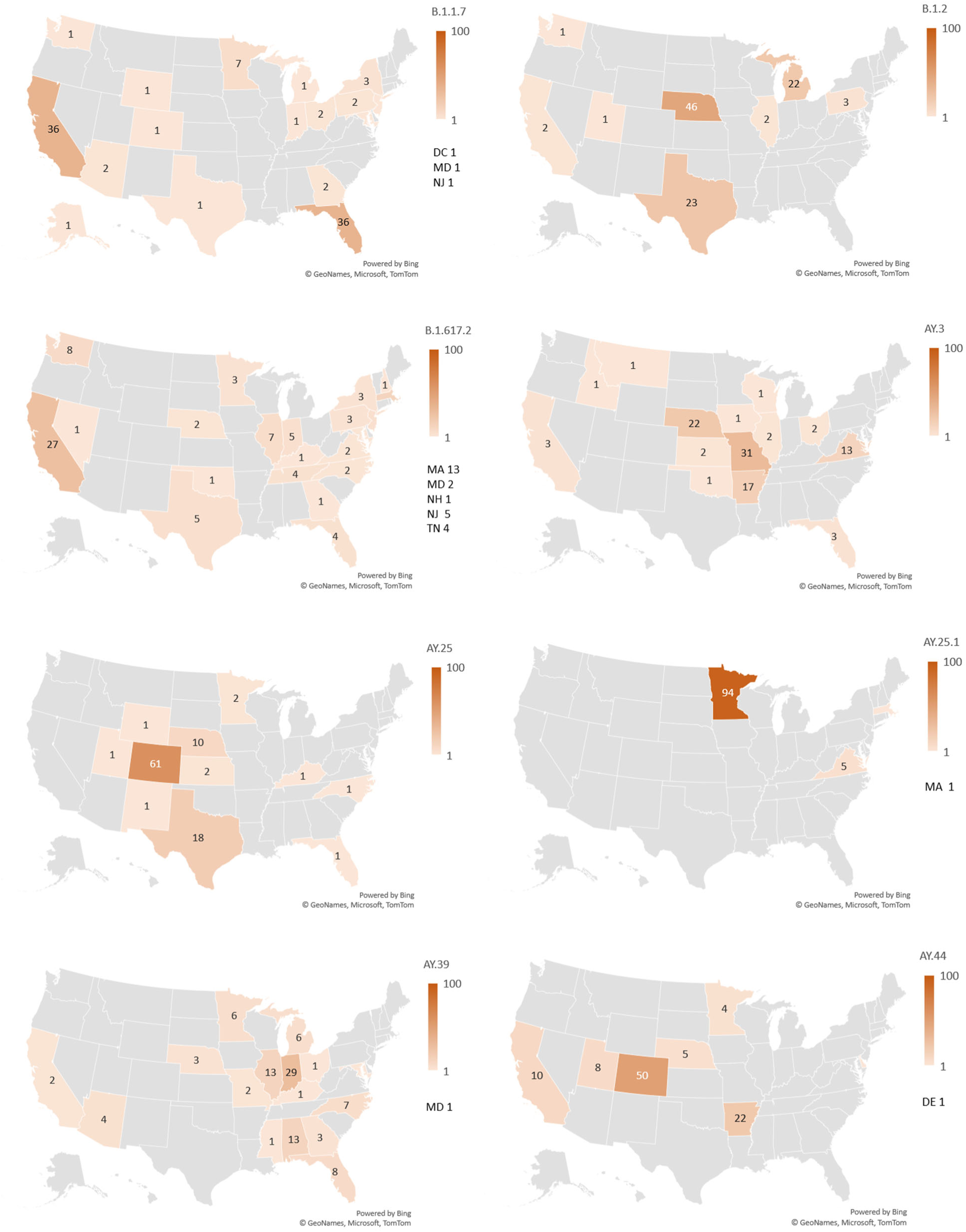
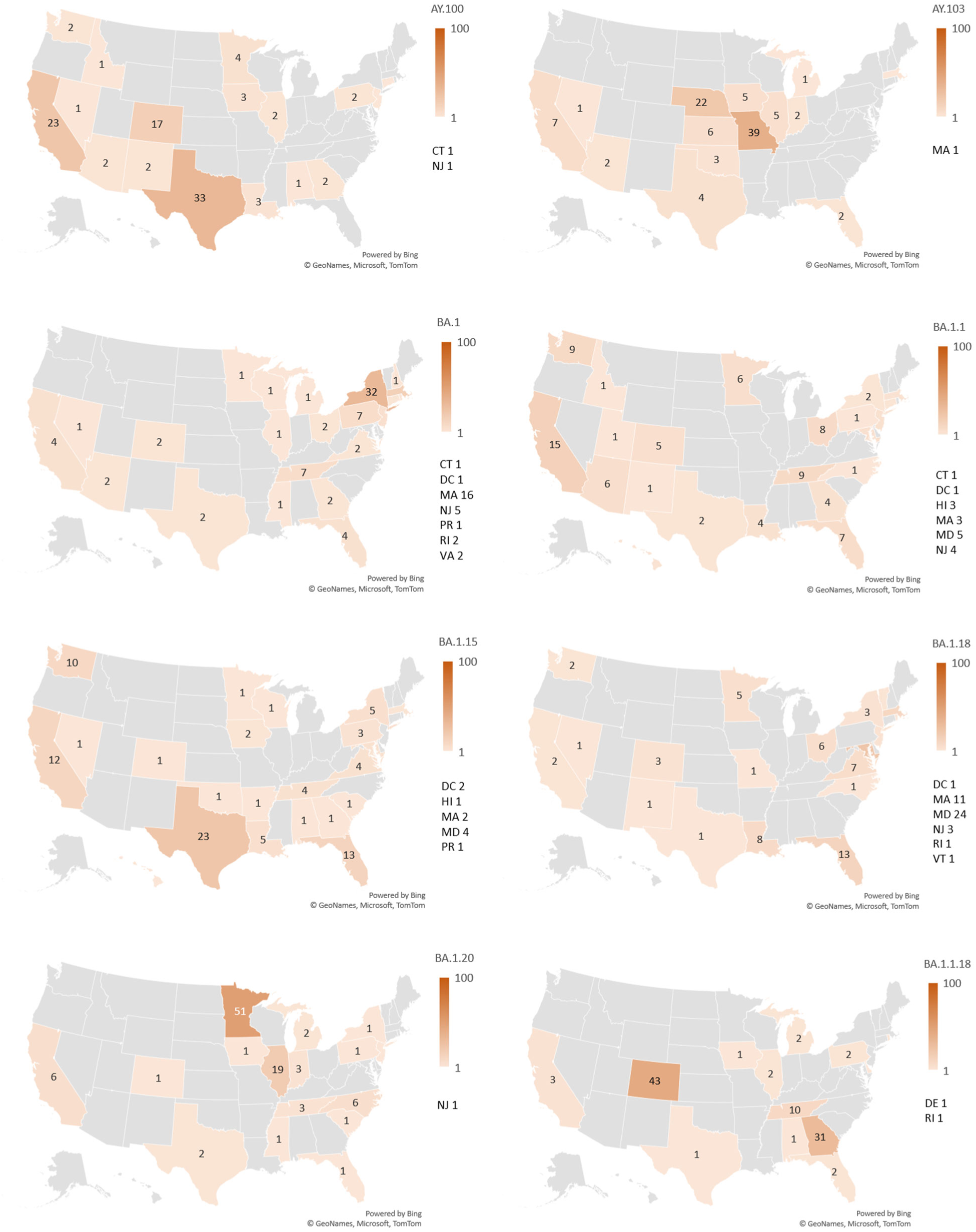
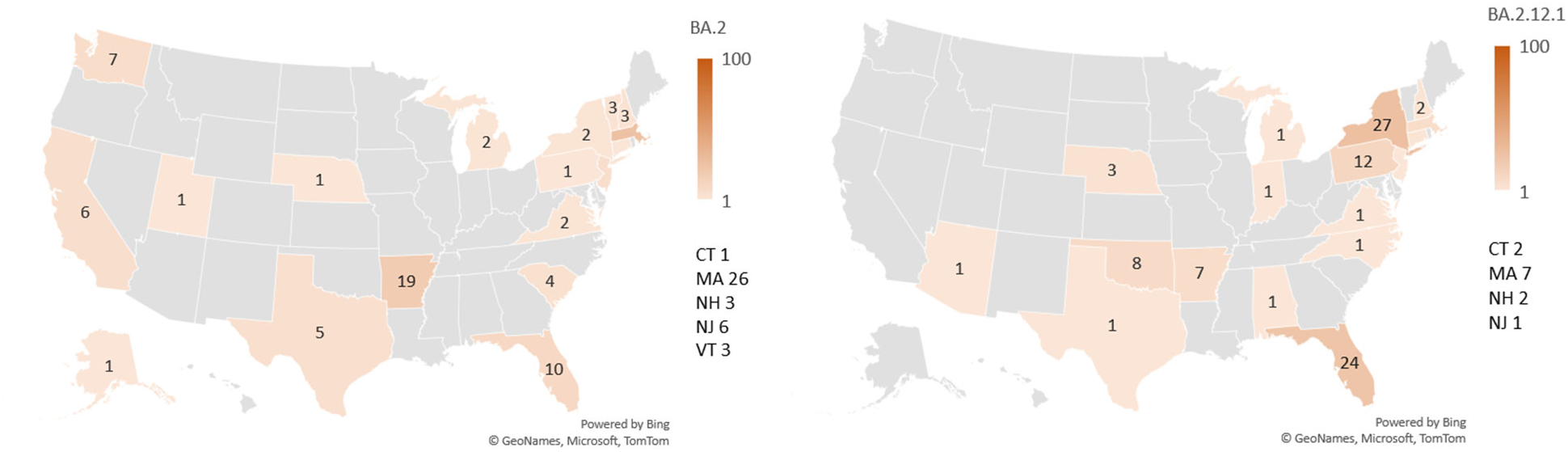
Heatmaps illustrating the spread of 18 SARS-CoV-2 variants across the US, showing the counts of the first 100 samples in each state for each variant.

**Table 1.**
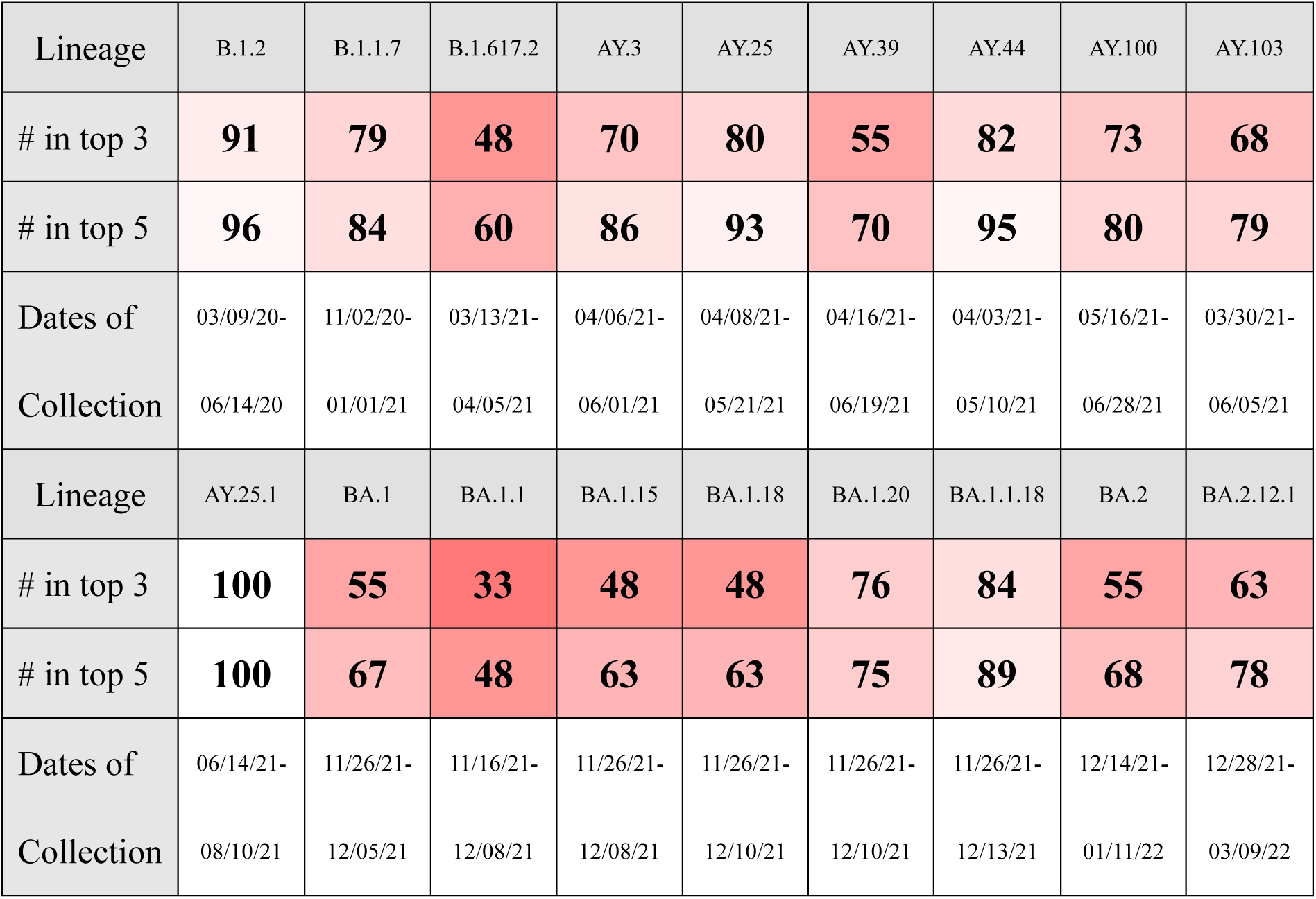
Total counts of samples collected in the top three and top five states in the US among the first 100 samples for 18 SARS-CoV-2 variants.

The extracted PRM patterns of Omicron BA.1.1 with more than 100 samples registered in GenBank are listed in Table 2, including the numbers of SF20 (NSF20) and ECDL (NECDL). Reversions of R346K were excluded from the analysis because it is regarded as a misregistration of a BA.1 variant. For comparison, Omicron BA.1.x variants in the control group are also listed in Table 2, with BA.1.1.7 and BA.1.1.15 excluded due to NECDL being smaller than 90. As this table shows, notably larger NSF20 values are observed in the PRMs of BA.1.1 compared to the control (BA.1.x) group.

**Table 2.** List of BA.1.1 PRMs with more than 100 entries and control BA.1 lineages, including the number of states where the first 20 samples were found (NSF20) and the number of entries with collection dates and locations (NECDL).

|  |  |  |  |  |  |  |  |
| --- | --- | --- | --- | --- | --- | --- | --- |
| Reversions at | 375 | 417-446 | 417-505 | 417-446, 764 | 417, 477-505 | 440 |  |
| NSF20 | **16** | **10** | **11** | **10** | **11** | **11** |  |
| NECDL | 123 | 778 | 119 | 129 | 333 | 232 |  |
| Reversions at | 440-446 | 446 | 477-505 | 493-505 | 547 | 856 | 417 |
| NSF20 | **12** | **17** | **9** | **7** | **14** | **14** | **4** |
| NECDL | 108 | 163 | 329 | 214 | 123 | 147 | 1012 |
| Lineage | BA.1.1.8 | BA.1.1.10 | BA.1.1.16 | BA.1.1.17 | BA.1.13 |  |  |
| NSF20 | **3** | **8** | **6** | **4** | **10** |  |  |
| NECDL | 734 | 507 | 611 | 299 | 113 |  |  |
| Lineage | BA.1.14 | BA.1.14.1 | BA.1.15.1 | BA.1.19 | BA.1.21 | BA.1.15.2 |  |
| NSF20 | **4** | **3** | **7** | **7** | **9** | **5** |  |
| NECDS | 245 | 97 | 598 | 693 | 102 | 1346 |  |

To assess whether there is a statistically significant difference in NSF20 between the PRM group and the control group, t-tests assuming unequal variance were conducted for all the listed data and the datasets with NECDL smaller than 1000. The obtained *p*-values in each case are 1.6 x 10^-4^ and 3.9 x 10^-5^ respectively, indicating that the PRMs were notably widespread from the beginning of their emergence.

Among the 13 kinds of PRMs listed in Table 2, six kinds had more than 50 data registered by December 27, 2021. The distribution of collection sites before that date is mapped in Figure 2. Since PRMs with reverse mutations from amino acids 417 to 446 in the spike protein exceeds 100 samples before December 27, 2021, the first 100 samples are mapped instead. Though some mutants have vague epicenters, others show scattered collection sites across the US, such as the PRM with reverse mutations at amino acids 417 and 477-505 in the spike protein.

**Figure 2.**
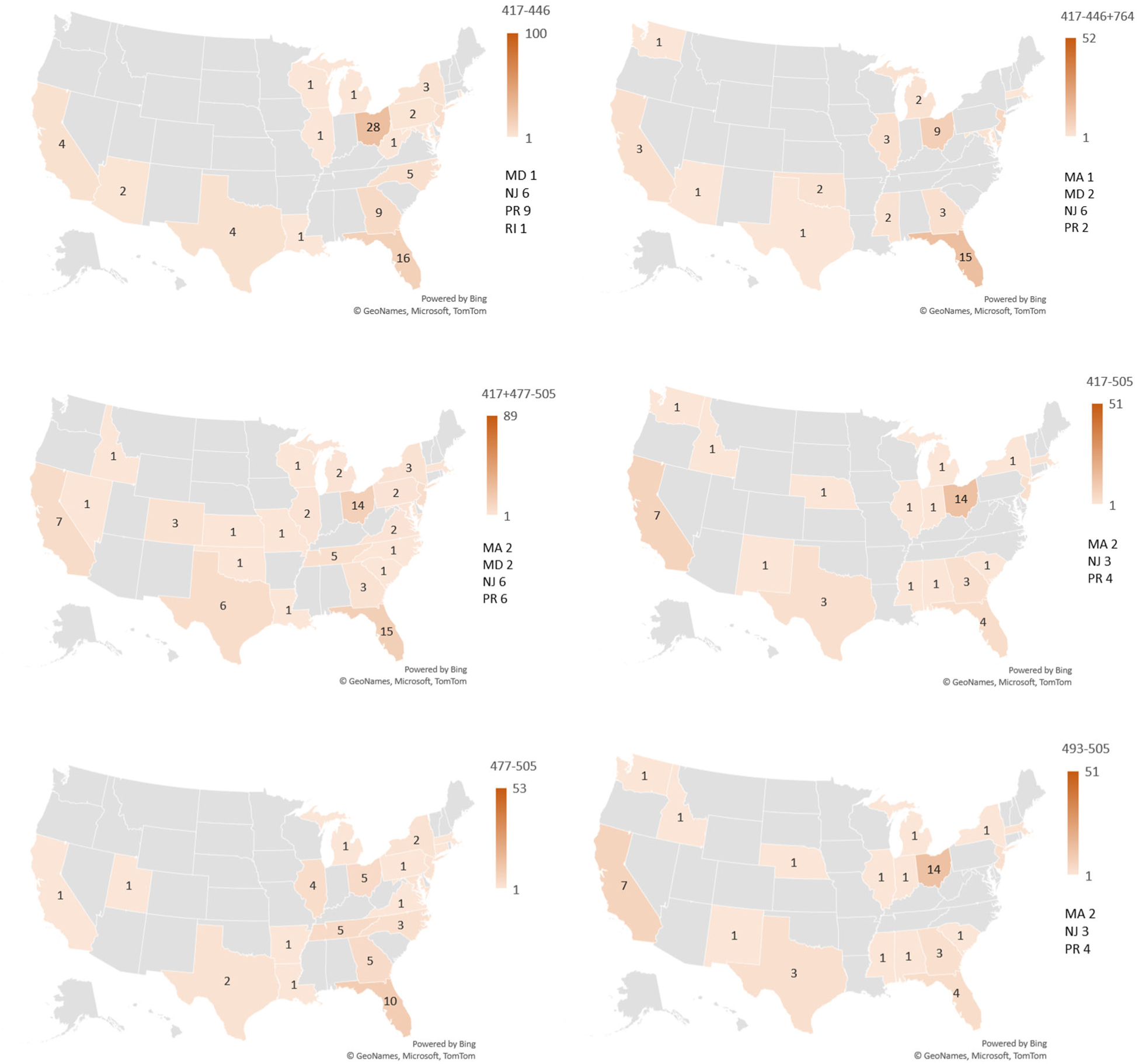
Heatmaps illustrating the spread of BA.1.1 PRMs across the US, showing the counts in each state by December 27, 2021. For PRMs with reversions at amino acids 417-446, the counts are based on the first 100 samples.

Chronological changes in the amount of collected sequences of BA.1 lineage and PRM sequences among them are shown in Figure 3, where data of BA.2 is also shown for comparison. What is significant is that the peaks of PRM collection in all variants appeared in the week starting December 24, 2021. The peak of the overall collection in BA.1, BA.1.15, BA.1.18, BA.1.20, and BA.20 synchronizes with the peak of PRM collection, while that in BA.1.1 and BA.1.1.18 appear three weeks later.

**Figure 3.**
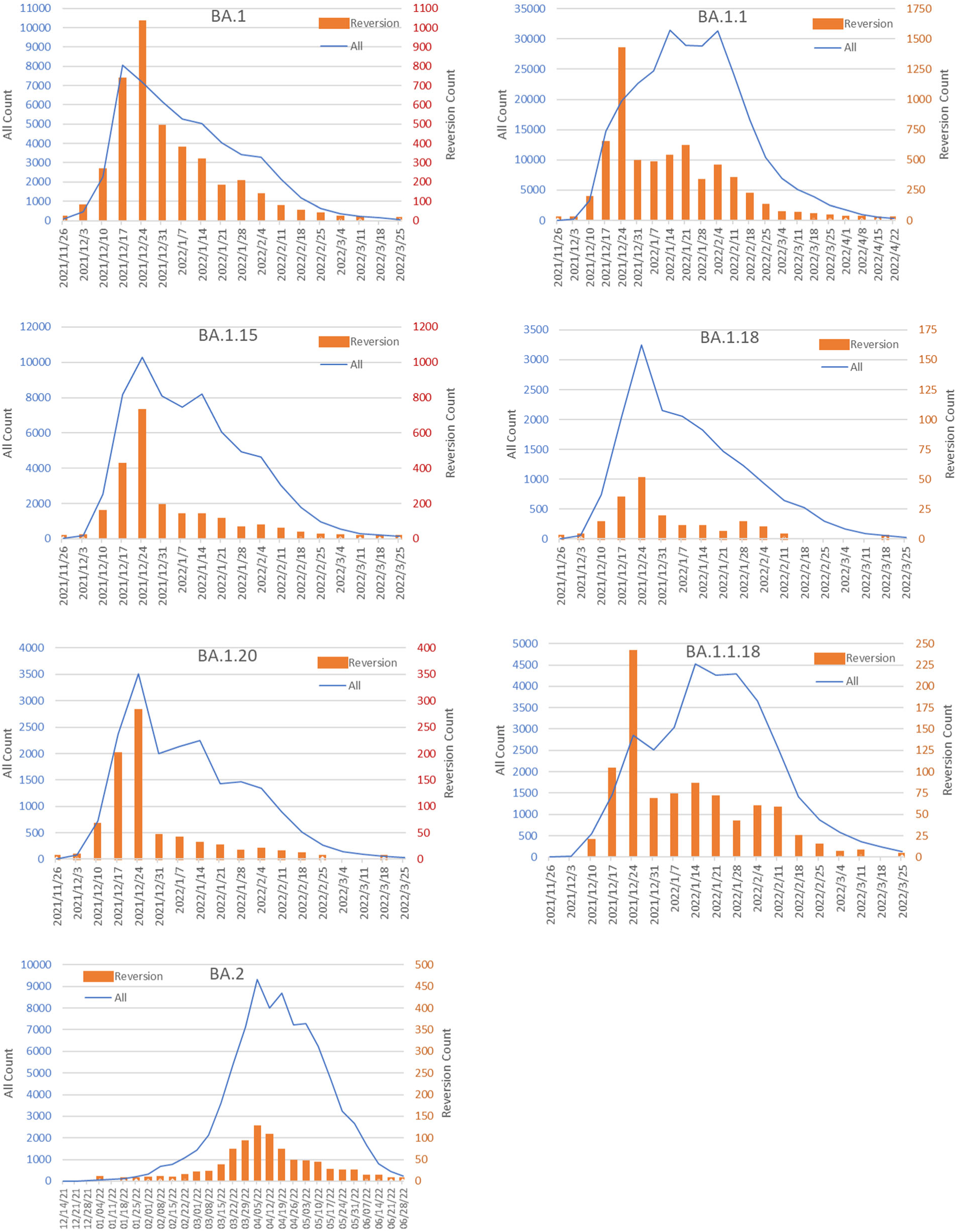
Histograms showing weekly collection counts in the US of each Omicron BA.1 and BA.2 variant and their PRMs. The horizontal axis shows the starting dates of each week for the counts.

To confirm data reliability, the rate of complete reads at Omicron mutation points in the spike protein registered to GISAID was calculated for Omicron BA.1 (which has the highest PRM rate), Omicron BA.1.1, and Omicron BA.1.18 (which has the lowest PRM rate), as shown in Figure 4 (A), where BA.1.18 has the highest rate and BA.1 has the lowest rate of complete reads. The ratio of organizations submitting ECDL to GenBank for each lineage is shown in Figure 4 (B), with the CDC Respiratory Viruses Branch, Division of Viral Diseases (CDC RVB) standing out for all variants.

**Figure 4.**
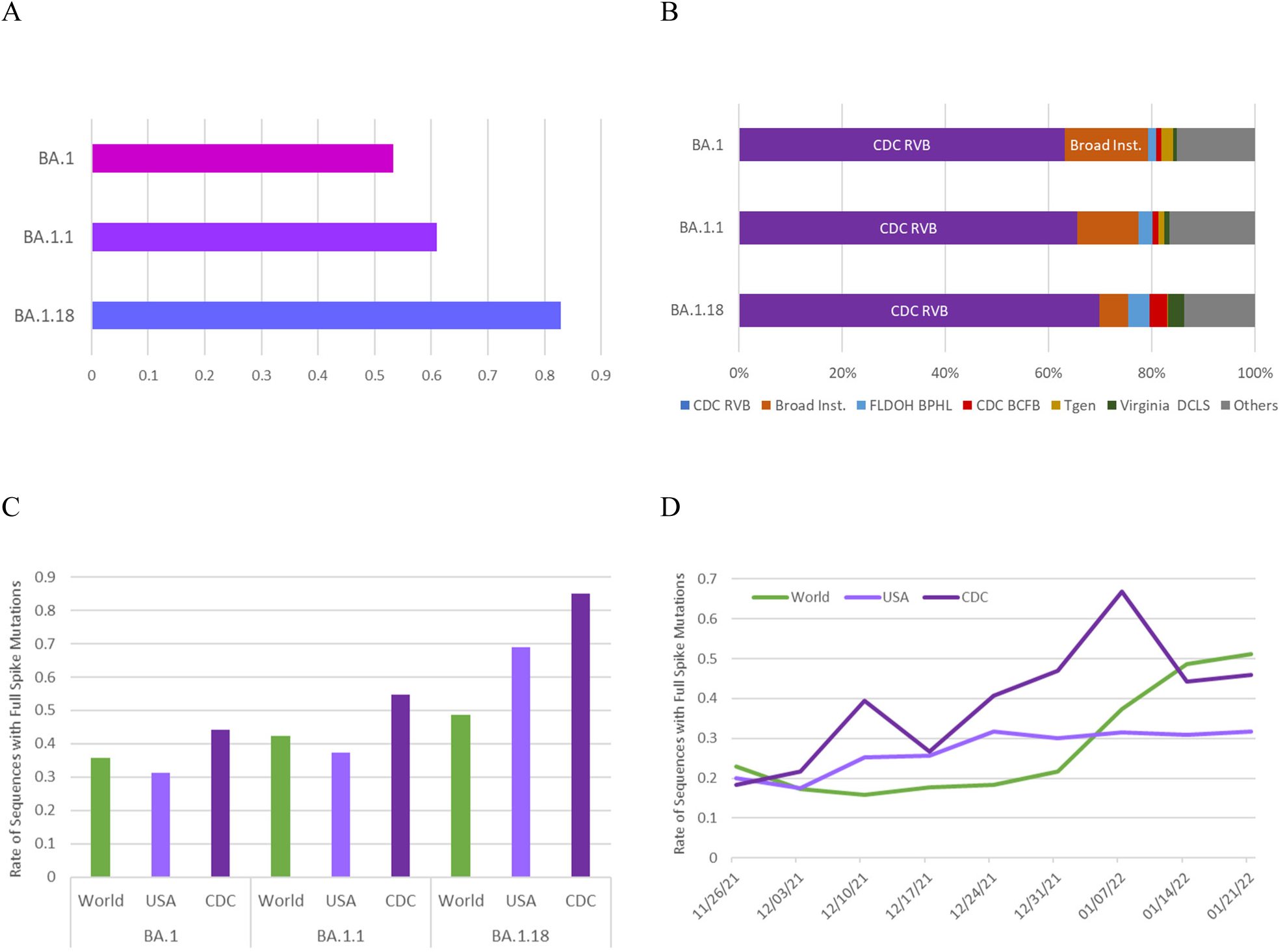
(A) The rate of complete reads at the Omicron mutation points of the spike protein registered to GISAID for BA.1, BA.1.1, and BA.1.18. (B) The ratio of organizations submitting ECDL to GenBank for BA.1, BA.1.1, and BA.1.18. (C) The ratio of EIASM specific to the consensus sequence of Omicron BA.1, registered to GISAID worldwide, in the US, and by the CDC. (D) Weekly ratios of EIASM-BA.1 registered to GISAID in the end of 2021 and the beginning of 2022. The horizontal axis shows the starting dates of the weekly counts.

Figure 4 (C) shows the ratio of EIASM-BA.1 registered to GISAID worldwide, in the US, and by the CDC. The read of the CDC has the highest ratio of EIASM for all variants, where BA.1.18 shows the highest and BA.1 shows the lowest ratio. Weekly ratios of EIASM registered to GISAID in the end of 2021 and the beginning of 2022 are shown in Figure 4 (D). The CDC has a higher ratio of EIASM from the onset of the Omicron surge.

Figure 5 presents the temporal changes in the number of collected PRM and all sequences registered to GenBank by organizations other than the CDC RVB, focusing on linages with a large number of registrations (BA.1, BA.1.1, BA.1.15, and BA.1.1.18). Compared with Figure 3, which includes data registered by the CDC RVB, a small shift in the peak is observed. However, the peak still synchronizes among all variants, and the PRM precedes the peak of all data for BA.1 and BA.1.1.18.

**Figure 5.**
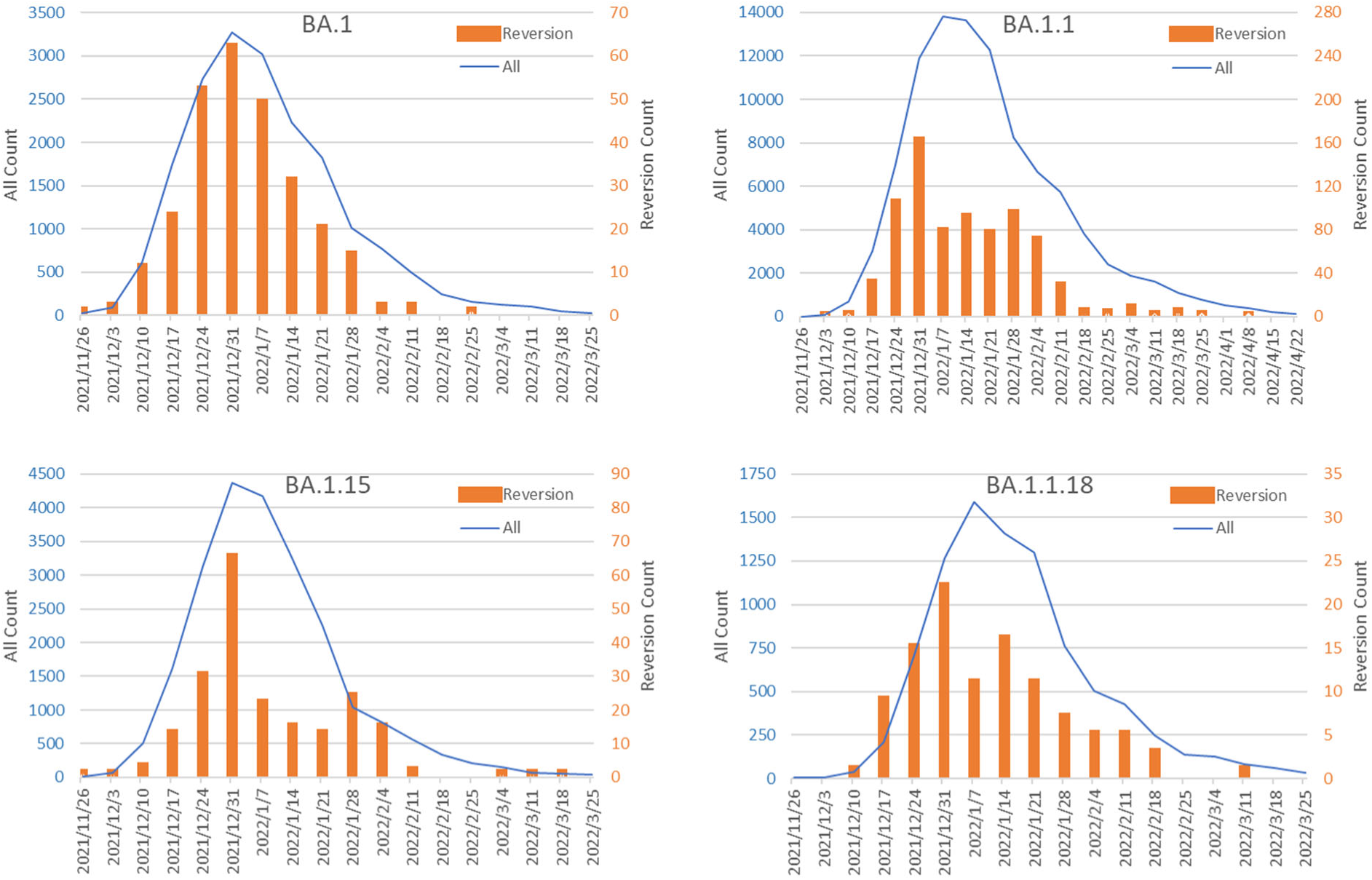
Histograms showing the weekly collection counts of each BA.1 variant and its PRM in the US, registered by organizations other than the CDC RVB. The horizontal axis shows the starting dates of the weekly counts.

## Discussion

The US-wide spread of Omicron BA.1.1 is notable among all the 18 variants analyzed in this study, showing a statistically significant difference from other variants in terms of epicenter ambiguity. While BA.1.1 is the most frequently registered variant in GenBank, with over 300,000 spike sequence entries, AY.44 and AY.103, ranking second and third in data size with over 200,000 entries, were not wide-spread from the beginning, with the former observed around Colorado and the latter around Missouri and Nebraska (Figure 1). It is also noteworthy that it took more than a month for AY.44 and AY.103 to reach 100 entries, while it took only about three weeks for BA.1.1 to reach 100 (Table 1). Excluding a single outlier sample collected on November 16, BA.1.1 sample collection began on November 26, making the time span for the first 100 sample collections less than two weeks.

Even among Omicron variants, BA.1.1, which has R346K point mutation in the spike protein, stands out. Other Omicron variants compared here have only a single mutation (NSP3_T1004I for BA.1.18, N_S33del for BA.1.20, NSP3_A1736V for BA.1.1.18) except for BA.1.15 (T10135C, NSP3_L106F, N_D343G). This suggests that even a small fraction of mutation cannot be synchronized all across the US, making the US-wide spread of BA.1.1 from the beginning of its emergence look all the more peculiar.

As described in the introduction, many studies have examined the epidemiology of Omicron variants all over the world, where the epicenters of transmission have generally been traceable. Therefore, the US-wide spread of Omicron BA.1.1 from the onset of its emergence, without clear epicenters, is extremely unusual. Any infection mechanisms currently known cannot account for the rapid US-wide transmission of the disease in such a short period.

The US-wide prevalence of Omicron BA.1.1 PRMs is also notable, showing a statistically significant difference from other BA.1 lineages that emerged during the same period of time (Table 2). This phenomenon was also confirmed with Omicron BA.1 in a previous study [40]. What is significant about Omicron BA.1.1 is that the surge of PRM sequences precedes that of all collected samples by a few weeks, which is observed only in Omicron BA.1.1 and not in BA.1 and BA.2 lineages (Figure 3).

Since PRM sequences and those in the control group were found within the same period of time and were genetically close, the difference observed cannot be attributed to seasonal or epidemiological factors. Notably, all kinds of PRMs disappeared quickly, suggesting they were not competitive against other Omicron BA.1 variants enough to spread widely through natural infections. Any theories of molecular biology, virology, or infectiology currently known cannot account for this sudden US-wide spread and wane of PRMs, either.

Some suggest that the Omicron variant may have originated from white-tailed deer. It is true that white-tailed deer were infected with SARS-CoV-2 and mutations occurred among them [41–43]. Though incubation in animal reservoirs can cause many mutations under completely different internal environment, synchronized animal-to-human transmission accompanied by the same mutational pattern all across the US is extremely unlikely to occur naturally.

One possible explanation for the anomalies observed in this paper is data errors. It is reported that Omicron variant sequences registered early were contaminated with Delta variant sequences due to primer issues, leading to massive appearance of reverse mutations [44]. Low ratios of EIASM in GISAID worldwide data during the early days of Omicron emergence supports this claim. Data registered by the CDC, however, shows higher EIASM ratios from the beginning (Figure 4D). The majority of US data registered in GenBank were submitted by the CDC (Figure 4B), which suggests that the data used in this study are reliable. Moreover, not only the synchronization of reverse mutations among the BA.1 lineage but also the peak of reverse mutation preceding that of all sequences in the BA.1.1 lineage is observed in the data registered by organizations other than the CDC RVB. This consistency also supports the reliability of the data used in this paper.

While the anomalies detected here can be explained by non-natural spread of infection, it is yet to be found what exactly happened during the early days of the Omicron surge in the US. Whether the phenomena discovered here resulted from natural or artificial causes, they should be taken seriously, as the Omicron variants have claimed a huge number of lives not only in the US [45] but also all around the world [46]. Further studies are needed to identify the cause of the anomalous US-wide transmission observed during the emergence of the Omicron variant.

## Data Availability

The source code used for this study is available at https://visual-media-lab.github.io/data/PRM_source_code/index.html

## References

[1] Pekar JE, Magee A, Parker E, et al. The molecular epidemiology of multiple zoonotic origins of SARS-CoV-2. Science 2022;377(6609):960–966. DOI:10.1126/science.abp8337

[2] Worobey M, Levy J, Serrano LM, et al. The Huanan Seafood Wholesale Market in Wuhan was the early epicenter of the COVID-19 pandemic. Science 2022;377(6609):951–959. DOI:10.1126/science.abp8715

[3] Massey SE, Jones A, Zhang D, et al. Unwarranted exclusion of intermediate lineage A-B SARS-CoV-2 genomes is inconsistent with the two-spillover hypothesis of the origin of COVID-19. Microbiology Research 2023;14(1):448–453. DOI: 10.3390/microbiolres14010033

[4] Daoyu Z, Gilles D, Adrian J, et al. Zoonosis at the Huanan Seafood Market: a critique. Zenodo 2022. DOI: 10.5281/zenodo.7169296

[5] Zhan SH, Deverman BE, Chan YA. SARS-CoV-2 is well adapted for humans. What does this mean for re-emergence? bioRxiv 2020. DOI: 10.1101/2020.05.01.073262

[6] Volz E, Hill V, McCrone JT, et al. (2021) Evaluating the effects of SARS-CoV-2 spike mutation D614G on transmissibility and pathogenicity, Cell 184(1), 64–75. DOI: 10.1016/j.cell.2020.11.020

[7] Korber B, Fischer WM, Gnanakaran S, et al. (2020) Tracking changes in SARS-CoV-2 spike: evidence that D614G increases infectivity of the COVID-19 virus. Cell 182(4), 812–827. DOI: 10.1016/j.cell.2020.06.043

[8] Hou YJ, Chiba S, Halfmann P, et al. (2020) SARS-CoV-2 D614G variant exhibits efficient replication ex vivo and transmission in vivo, Science 370(6523), 1464–1468 DOI: 10.1126/science.abe8499

[9] Du Plessis L, McCrone JT, Zarebski AE, et al. Establishment and lineage dynamics of the SARS-CoV-2 epidemic in the UK. Science 2021;371:708–712 DOI: 10.1126/science.abf2946

[10] Miyata T, Yasunaga T. Molecular evolution of mRNA: A method for estimating evolutionary rates of synonymous and amino acid substitutions from homologous nucleotide sequences and its application. J Molecular Evolution 1980;16(1):23–36. DOI: 10.1007/BF01732067

[11] Li WH, Wu CI, Luo CC. A new method for estimating synonymous and nonsynonymous rates of nucleotide substitution considering the relative likelihood of nucleotide and codon changes. Molecular Biology and Evolution 1985;2(2):150–174. DOI: 10.1093/oxfordjournals.molbev.a040343

[12] Nikolaidis M, Papakyriakou A, Chlichlia K, et al. Comparative analysis of SARS-CoV-2 variants of concern, including Omicron, highlights their common and distinctive amino acid substitution patterns, especially at the spike ORF. Viruses 2022;14(4):707. DOI: 10.3390/v14040707

[13] Hassan SS, Kodakandla V, Redwan EM, et al. Non-uniform aspects of the SARS-CoV-2 intraspecies evolution reopen question of its origin. International Journal of Biological Macromolecules 2022; 222:972–993. DOI: 10.1016/j.ijbiomac.2022.09.184

[14] Callaway E. Heavily mutated Omicron variant puts scientists on alert. Nature 2021;600(7887):21. DOI: 10.1038/d41586-021-03552-w

[15] Jung C, Kmiec, D, Koepke L, et al. Omicron: what makes the latest SARS-CoV-2 variant of concern so concerning? J Virology 2022;96(6):e02077–21. DOI: 10.1128/jvi.02077-21

[16] Mallapaty C. The hunt for the origin of Omicron. Nature 2022;602(7898):26–28. DOI: 10.1038/d41586-022-00215-2

[17] Choi B, Choudhary MC, Regan J, et al. persistence and evolution of SARS-CoV-2 in an immunocompromised host. The New England Journal of Medicine 2020;383(23):2291–2293. DOI: 10.1056/NEJMc2031364

[18] Kemp SA, Collier DA, Datier RP, et al. SARS-CoV-2 evolution during treatment of chronic infection. Nature 2021;592:277–282. DOI: 10.1038/s41586-021-03291-y

[19] Truong TT, Ryutov A, Pandey U, et al. Increased viral variants in children and young adults with impaired humoral immunity and persistent SARS-CoV-2 infection: A consecutive case series. EBioMedicine 2021 May;67:103355. DOI: 10.1016/j.ebiom.2021.103355

[20] Wei C, Shan KJ, Wang W, et al. Evidence for a mouse origin of the SARS-CoV-2 Omicron variant. J Genet Genomics 2021;48(12):1111–1121. DOI: 10.1016/j.jgg.2021.12.003

[21] Zhang W, Shi K, Geng Q, et al. Structural basis for mouse receptor recognition by SARS-CoV-2 omicron variant. PNAS 2022; 119(44): e2206509119. DOI: 10.1073/pnas.2206509119

[22] Piplani S, Singh PK, Winkler DA, et al. In silico comparison of SARS-CoV-2 spike protein-ACE2 binding affinities across species and implications for virus origin. Scientific Reports 2021;11:13063. DOI: 10.1038/s41598-021-92388-5

[23] Kakeya H, Matsumoto Y. A probabilistic approach to evaluate the likelihood of artificial genetic modification and its application to SARS-CoV-2 Omicron variant. ISPJ Trans Bioinformatics 2022;15:22–29. DOI: 10.2197/ipsjtbio.15.22

[24] Kakeya H, Arakawa H, Matsumoto Y. Multiple probabilistic analyses suggest non-natural origin of SARS- CoV-2 Omicron variant. Zenodo 2023. DOI: 10.5281/zenodo.7470652

[25] Tanaka A, Miyazawa T. Unnaturalness in the evolution process of the SARS-CoV-2 variants and the possibility of deliberate natural selection. Zenodo 2023. DOI: 10.5281/zenodo.8361577

[26] Kakeya H, Matsumoto Y. Repeated emergence of probabilistically and chronologically anomalous mutations in SARS-CoV-2 during the COVID-19 pandemic. Zenodo 2023. DOI: 10.5281/zenodo.8216232

[27] Kakeya H, Kanazaki T. Anomalous biases of reverse mutations in SARS-CoV-2 variants. Jxiv 2023. DOI: 10.51094/jxiv.545

[28] Mella-Torres A, Escobar A, Barrera-Avalos C, et al. Epidemiological characteristics of Omicron and Delta SARS-CoV-2 variant infection in Santiago, Chile. Front Public of Health 2022;10:98443 DOI:10.3389/fpubh.2022.984433

[29] Liu L, Chiou S, Chen P, et al. Epidemiology and analysis of SARS-CoV-2 Omicron subvariants BA.1 and 2 in Taiwan. Scientific Reports 2023; 13:16583 DOI: 10.1038/s41598-023-43357-7

[30] Lamarca AP, Souza, U.J.B.d., Moreira, F.R.R., et al. The Omicron Lineages BA.1 andBA.2 (Betacoronavirus SARS-CoV-2) Have Repeatedly Entered Brazil through a Single Dispersal Hub. Viruses 2023;15:888 DOI:10.3390/v15040888

[31] Freitas MTS, Sena LOC, Fukutani KF, et al. The increase in SARS-CoV-2 lineages during 2020–2022 in a state in the Brazilian Northeast is associated with a number of cases. Front Public of Health 2023; DOI:10.3389/fpubh.2023.1222152

[32] Rodrigues ES, Slavov SN, de La Roque D.G.L, et al. Epidemiology of the SARS-CoV-2 Omicron Variant Emergence in the Southeast Brazilian Population. Microorganisms 2024;12:449. DOI:10.3390/microorganisms1230449

[33] Zárate S, Taboada B, Rosales-Rivera M, et al. Omicron-BA.1 Dispersion Rates in Mexico Varied According to the Regional Epidemic Patterns and the Diversity of Local Delta Subvariants. Viruses 2023;15: 243. DOI:10.3390/v15010243

[34] Tsui J, McCrone JT, Lambert B, et al. Genomic assessment of invasion dynamics of SARS-CoV-2 Omicron BA.1. Science 2023;381:336–343 DOI: 10.1126/science.adg6605

[35] Elliott P, Eales O, Steyn N, et al. Twin peaks: The Omicron SARS-CoV-2 BA.1 and BA.2 epidemics in England. Science 2022;376:1432 DOI: 10.1126/science.abq4411

[36] Tong C, Shi W, Siu GH, et al. Understanding spatiotemporal symptom onset risk of Omicron BA.1, BA.2 and hamster-related Delta AY.127. Frontiers in Public Health2022; 10:978052 DOI: 10.3389/fpubh.2022.978052

[37] Tegally H, Moir M, Everatt J, et al. Emergence of SARS-CoV-2 Omicron lineages BA.4 and BA.5 in South Africa. Nature Medicine 2022;28:1785-1790 DOI: 10.1038/s41591-022-01911-2.

[38] Chrysostomou AC, Vrancken B, Haralambous C, et al. Unraveling the Dynamics of Omicron (BA.1, BA.2, and BA.5) Waves and Emergence of the Deltacron Variant: Genomic Epidemiology of the SARS-CoV-2 Epidemic in Cyprus (Oct 2021–Oct 2022). Viruses 2023;15:1933 DOI: 10.3390/v15091933

[39] Lopes L, Pham K, Klaassen F, et al. Combining genomic data and infection estimates to characterize the complex dynamics of SARS-CoV-2 Omicron variants in the US. Cell Reports 2024; 43:114451 DOI:10.1016/j.celrep.2024.114451

[40] Kakeya H. Anomalous US-wide prevalence of reversion mutants in the emergence of Omicron BA.1. Zenodo 2024. https://zenodo.org/records/13158254

[41] Pickering B, Lung O, Maguire F, et al. Divergent SARS-CoV-2 variant emerges in white-tailed deer with deer-to-human transmission. Nature Microbiology 2022; 7:2011–2024 DOI:10.1038/s41564-022-01268-9

[42] Feng A, Bevins S, Chandler J, et al. Transmission of SARS-CoV-2 in free-ranging white-tailed deer in the United States. Nature Communications 2023; 14:4078 DOI:10.1038/s41467-023-39782-x

[43] McBride DS, Garushyants SK, Franks J, et al. Accelerated evolution of SARS-CoV-2 in free-ranging white-tailed deer. Nature Communications 2023; 14:5105 DOI:10.1038/s41467-023-40706-y

[44] Martin DP, Lytras S, Lucaci AG, et al. Selection analysis identifies clusters of unusual mutational changes in Omicron lineage BA.1 that likely impact spike function. Mol Biol Evol 2022;39(4):msac061. DOI: 10.1093/molbev/msac061

[45] Paglino E, LundBerg DJ, Zhou Z, et al. Monthly excess mortality across counties in the United States during the COVID-19 pandemic, March 2020 to February 2022. Sci ADV 2023;9:eadf9742. DOI: 10.1126/sciadv.adf9742

[46] Mostert S, Hoogland M, Huibers, M, et al. Excess mortality across countries in the Western World since the COVID-19 pandemic: ‘Our World in Data’ estimates of January 2020 to December 2022. BMJ Public Health 2024;2:e000282. DOI: 10.1136/bmjph-2023-000282

